# The Oregon Child Absenteeism Due to Respiratory Disease Study (ORCHARDS): Rationale, Objectives, and Design

**DOI:** 10.1101/2021.02.01.21250878

**Authors:** Jonathan L. Temte, Shari Barlow, Maureen Goss, Emily Temte, Amber Schemmel, Bradley Maerz, Cristalyne Bell, Lily Comp, Mitchell Arnold, Kimberly Breunig, Sarah Clifford, Erik Reisdorf, Peter Shult, Mary Wedig, Thomas Haupt, James Conway, Ronald Gangnon, Ashley Fowlkes, Amra Uzicanin

## Abstract

**Background:** Influenza viruses pose significant disease burdens through annual seasonal outbreaks and unpredictable pandemics. Existing influenza surveillance programs have relied heavily on reporting of medically attended influenza (MAI). Continuously monitoring cause-specific school absenteeism may identify local acceleration of seasonal influenza activity. The Oregon Child Absenteeism Due to Respiratory Disease Study (ORCHARDS; Oregon, WI) implements daily school-based monitoring of influenza-like illness–specific student absenteeism (a-ILI) in pre-kindergarten through grade 12 schools and assesses this approach for early detection of accelerated influenza and other respiratory pathogen transmission in schools and surrounding communities.

**Methods:** Starting in September 2014, ORCHARDS combined automated reporting of daily absenteeism within 6 schools and home visits to school children with acute respiratory infections (ARI). Demographic, epidemiological, and symptom data are collected along with respiratory specimens. Specimens are tested for influenza and other respiratory viruses. Household members can opt into a supplementary household transmission study. Community comparisons are possible using a pre-existing, long-standing, and highly effective influenza surveillance program, based on MAI at 5 primary care clinics in the same geographical area.

**Results:** Over the first 5 years, a-ILI occurred on 6,634 (0.20%) of 3,260,461 student school days. Viral pathogens were detected in 64.5% of the 1,728 children with ARI who received a home visit. Influenza was the most commonly detected virus, noted in 23.3% of ill students. Influenza (p<0.001) and adenovirus (P=0.004) were significantly associated with a-ILI.

**Conclusion:** ORCHARDS uses a community-based design to detect influenza trends over multiple seasons and to evaluate the utility of absenteeism for early detection of accelerated influenza and other respiratory pathogen transmission in schools and surrounding communities. Initial findings suggest the study design is succeeding in collecting appropriate data to achieve study objectives.

## I. Background

Influenza viruses pose significant disease burdens through annual seasonal outbreaks and unpredictable pandemics. Seasonal outbreaks result in an average annual toll of nearly 12,000 to 61,000 deaths in the United States (1) and are associated with an estimated annual cost of $11.2 billion.(2) While infrequent—only 4 have occurred since 1918—influenza pandemics are characterized by large numbers of cases and increased morbidity and mortality.(3) Interventions for seasonal and pandemic influenza include vaccines, antiviral medications, and nonpharmaceutical interventions (NPI), also known as community mitigation, but their combined success is predicated on timely deployment. NPI, the first line of defense (4), hinges on early detection and recognition of outbreaks.(4-6)

Existing influenza surveillance programs rely on ambulatory and inpatient medical facilities to report cases of influenza-like illness (ILI) and test-confirmed influenza.(7,8) Even though influenza transmission among school-aged children frequently precedes subsequent community transmission (9,10), there has been no effort to systematically evaluate whether school-based monitoring of influenza activity can complement routine surveillance or even serve as an early-warning system for increased influenza activity in schools and the wider community. Monitoring of school absenteeism is feasible, as seasonal outbreaks generally occur between late fall and early to mid-spring while schools are in session (11,12), and most of the 13,588 school districts (13) across the United States routinely collect daily absenteeism data using electronic school information systems.(14)

Early evaluations have shown promise. During the 2009 influenza pandemic, there was high correlation (r=0.92) between hospitalized influenza cases and school absenteeism due to ILI.(15) This strong association, however, was likely due to the short and intense nature of the outbreak, which amplified absenteeism related to ILI. Conversely, monitoring all-cause absenteeism was less effective due to the “noise” associated with school data and the multi-factorial nature of absenteeism.(16-18) The value of continuously monitoring cause-specific absenteeism, such as ILI, over the entire school year to identify local activity acceleration of seasonal influenza is not well researched or understood.

## II. ORCHARDS overview statement

The goal of Oregon Child Absenteeism Due to Respiratory Disease Study (ORCHARDS) is to develop and implement a system for daily school-based monitoring of ILI-specific student absenteeism in pre-kindergarten (4K) through grade 12 schools and assess the system’s usability for early detection of accelerated influenza and other respiratory pathogen transmission in schools and surrounding communities. The theoretical relationships between influenza infection in school-aged children, K-12 absenteeism, and influenza infection in the surrounding community are demonstrated in Figure 1.

**Figure 1.**
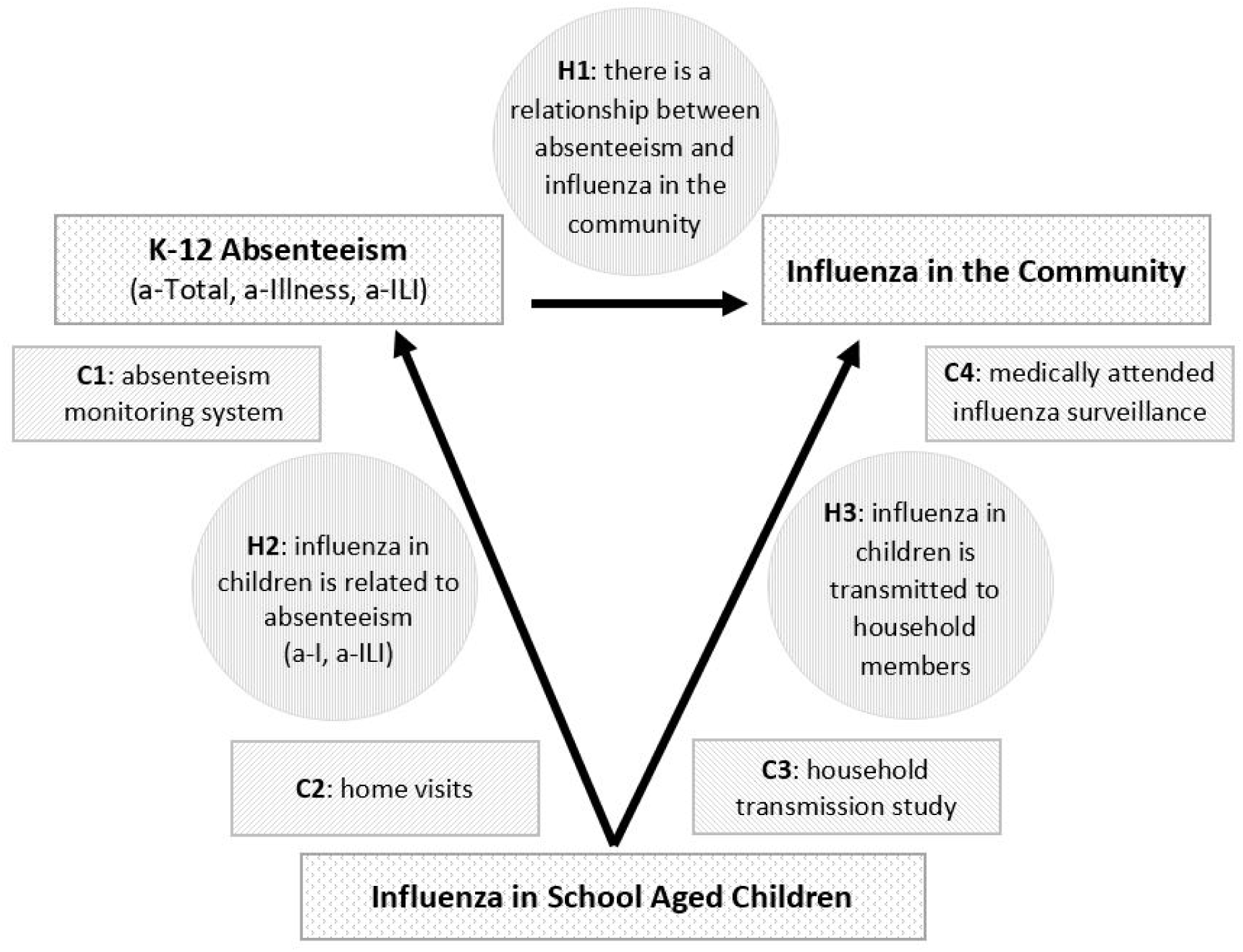
Theoretical framework of ORCHARDS demonstrating the relationships between influenza in school-aged children, K-12 school absenteeism, and medically attended influenza in the community. The relatedness of the four components (C1-C4) of ORCHARDS and the three primary hypotheses (H1-H3) are provided.

## III. Specific objectives and study hypotheses

### ORCHARDS objectives

- Develop an automated cause-specific school absentee monitoring system to identify school absences related to ILI in 4K-12 students across the selected school district on a daily basis (Figure 1; component 1).
- Determine the etiology of ILI in absent students (Figure 1; component 2).
- Detect within-household transmission of influenza in households from which a student has been absent from school due to ILI (Figure 1; component 3).
- Compare the data from ILI- and influenza-specific student absenteeism in the participating schools to data routinely collected in complementary influenza surveillance in the health care facilities serving the same general population of the school district (Figure 1; component 4).

### Study hypotheses

- There is a relationship between illness-specific absenteeism and influenza infection in the community (Figure 1; hypothesis 1).
- Influenza infection in school-aged children is related to absenteeism (Figure 1; hypothesis 2).
- Influenza infection in school-aged children is transmitted to household members (Figure 1; hypothesis 3).

## IV. Methods

### IVa. Location and timeframe

#### Location

The Oregon School District (OSD: www.oregonsd.org) encompasses the villages of Oregon and Brooklyn, which are located in rural areas of Dane County, Wisconsin and span approximately 85 square miles (figure 2. Lat: 42.90 N, Long: 89.43W; altitude: 1,053 feet).(19) The ORCHARDS team selected OSD based on school population size, familiarity, proximity to study resources, and coverage by an existing and successful MAI surveillance program. The region experiences distinct temperate seasonality with significant variations in temperature, precipitation, and humidity.

**Figure 2.**
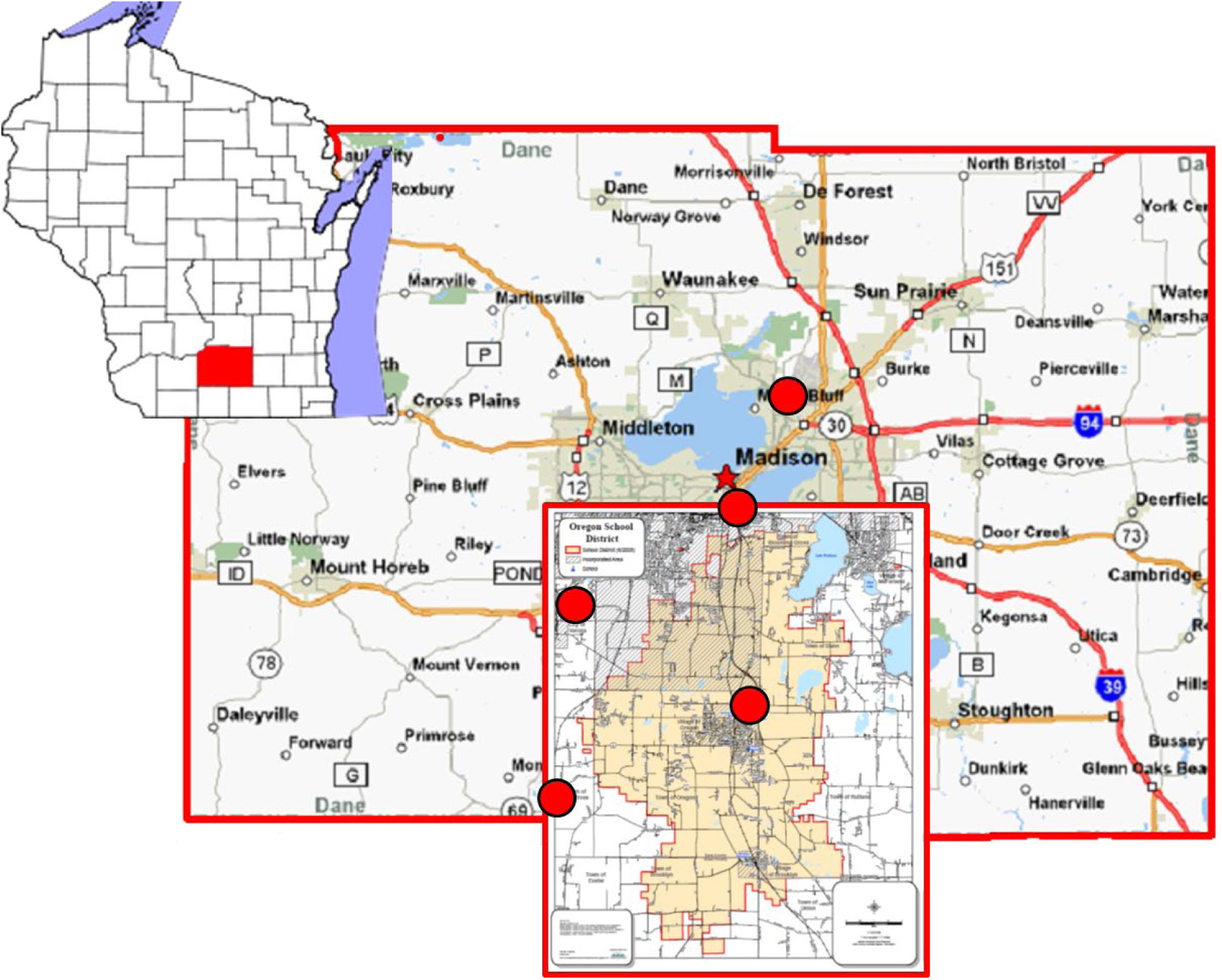
Location of the Oregon School District (OSD) in the Villages of Oregon and Brooklyn in southcentral Wisconsin. OSD extend past the southern border of Dane County and is drawn to scale. The red dots indicate locations of family medicine clinics that have participated in long-term surveillance of medically attended influenza. ^*^Wisconsin state image created by David Benbennick Feb. 13, 2006. Image released to public domain: https://en.wikipedia.org/wiki/Dane_County,_Wisconsin#/media/File:Map_of_Wisconsin_highlighting_Dane_County.svg ^†^ Dane County image was adapted from a Wisconsin Department of Transportation map that is licensed for non-commercial use and is in the public domain: http://lis202sec308.pbworks.com/w/page/91834941/DaneCounty%2C ^‡^ The Oregon School District map was obtained from the district with permission to modify. The original graphic has been updated and is in the public domain: https://www.oregonsd.org/Page/5396

#### Community

The OSD population is estimated at 20,094 and is less racially and ethnically diverse, wealthier, and better educated than the average community in the United States (Table 1) (20). Ages of individual family members and mean household size are similar to the US average. The population is mainly distributed in the villages of Oregon (10,390) and Brooklyn (1,255) with the remainder in decentralized subdivisions and farms.

**Table 1.**
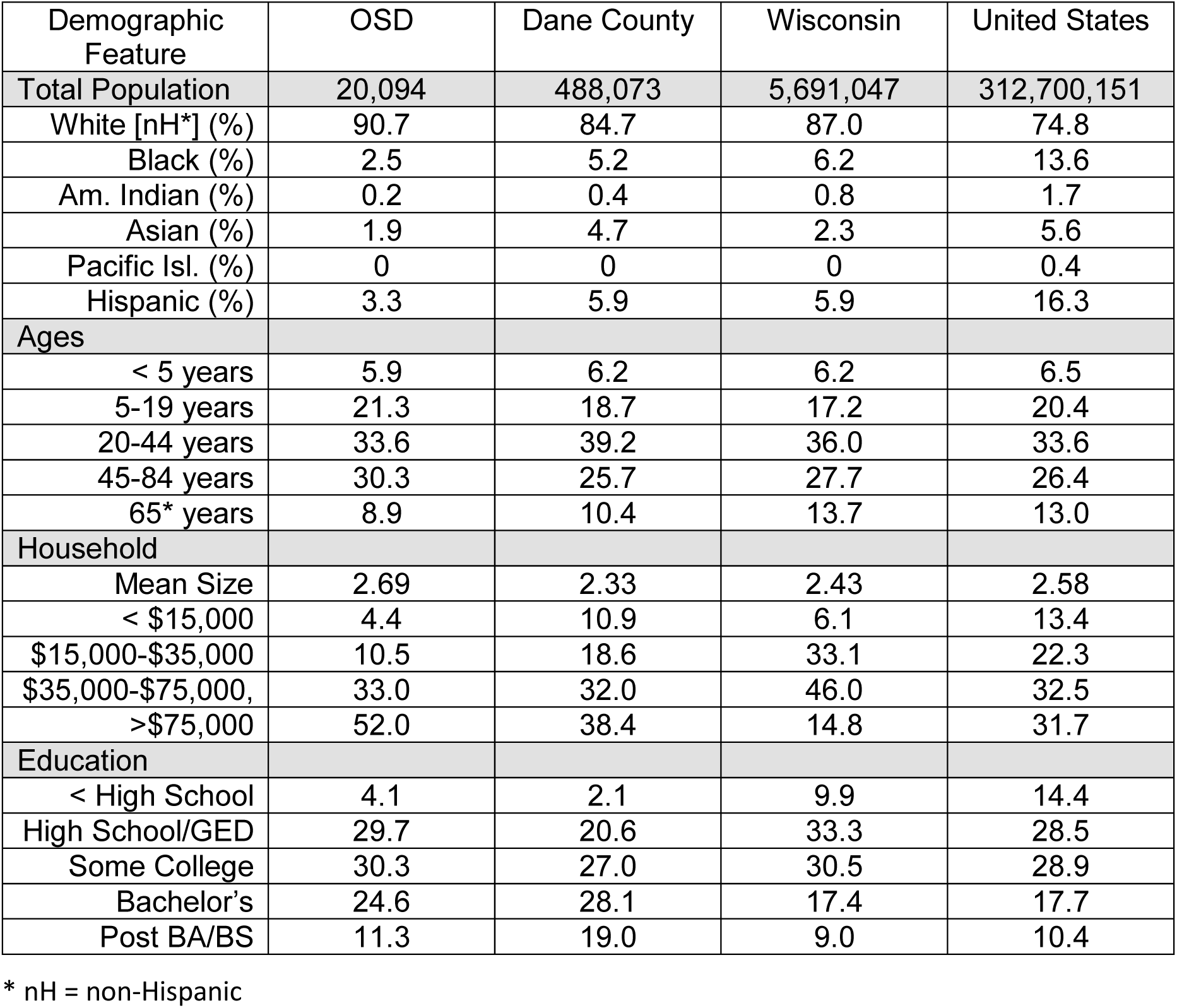
Demographic descriptions of the Oregon School District (OSD) as compared to Dane County, Wisconsin, and the United States. Data from US Census.

The OSD is comprised of 6 public schools with a growing enrollment, estimated at 4,091 students (19% of total population) during the 2018-2019 school year.(21) There are 3 elementary schools (grade K-4)— 2 in Oregon and 1 in Brooklyn—with a combined 1,503 students. There is one intermediate school (grade 5-6) with 623 students and one middle school (grade 7-8) with 596 students, both located in Oregon. The high school (grade 9-12) has 1,145 students and is located in Oregon. There are 224 children enrolled in 4K programs.

#### Timeframe

Initiation of ORCHARDS data collection occurred in phases. Absenteeism data collection (objective 1) commenced at the beginning of the 2014-2015 academic year on September 2, 2014. Data collection from student home visits (objective 2) commenced January 5, 2015, immediately following the winter break. The household transmission sub-study (objective 3) started September 6, 2016. MAI surveillance (objective 4) has been ongoing since October 2009.(8) Whereas absenteeism data do not accrue during planned and unplanned school breaks—including summer and weekends—home visits with ill students and the household transmission study continue throughout the year. All components of ORCHARDS have continued without interruption until present.

### IVb. Absenteeism monitoring system

#### Reporting unscheduled absences

We introduced minimal modifications to an existing absenteeism reporting system. Parents report unscheduled absences to attendance staff using an automated telephone system. Callers are prompted to provide the student’s name and the reason for absence, including symptoms if the child has a cold or flu-like illness. We worked with OSD to provide uniform messaging on each school’s absentee line with additional information pertaining to influenza-like illness (ILI) symptoms:

> *“Please inform us if your child has any flu-like symptoms such as fever with cough, sneezing, chills, sore throat, body aches, fatigue, runny nose, and/or stuffy nose*.*”*

In the event that a student is absent without a report, OSD attendance staff—in the interest of child safety—make repeated efforts to contact the home or parents/guardians before the end of the day.

#### Absenteeism definition

Because of variability among the OSD schools in terms of the number of class periods, from 2 to 10 per day, for which a child can be absent and for simplicity/generalizability of electronic data retrieval, if a student is absent part of a school day, we consider them absent for the entire day.

#### Types of absenteeism

All-cause or total absenteeism (a-TOT) is defined as an absence for any reason. Absence due to illness (a-I) is an absence due to any reported illness. Absence due to ILI (a-ILI) is a subset of a-I for which ILI symptoms are reported. These operational definitions likely underestimated a-I and a-ILI due to under-reporting by parents.

#### a-ILI definition

We considered established definitions for ILI.(22-23) For simplicity of recognition by non-medical attendance staff members, we used a simplified version of the CDC standard definition. ILI for ORCHARDS is defined as the presence of fever and at least one respiratory tract symptom (cough, sore throat, nasal congestion, or runny nose). We selected these symptoms based on data from the Wisconsin component of the Influenza Incidence Surveillance Project (W-IISP).(8) Reported fever plus one or more of these symptoms, as compared to respiratory symptoms in the absence of fever, is associated with a 9-fold increase in underlying influenza infection in school-aged children presenting for primary medical care (Temte, unpublished).

#### Data system

OSD utilizes Infinite Campus^®^ (24), a commercially available electronic student information system (SIS), to track student scheduling, enrollment, performance, and attendance. This system allows attendance staff to identify a student, select a period, and select a reason for absence from a modifiable, drop-down pick list. The OSD Information Technology (IT) staff added an option for “Absent due to influenza-like illness” (a-ILI); this required less than 5 minutes to enable.

#### Data extraction and secure file transfer

OSD IT staff created an automated process within Infinite Campus^®^ to extract daily counts of absent individuals by school, grade, and absence type (a-TOT, a-I and a-ILI). The data contain no personal identifiers and are fully compliant with the Family Educational Rights and Privacy Act (FERPA: 20 U.S.C. § 1232g; 34 CFR Part 99).(25) The data are sent to a secure file transfer protocol (ftp) site at the University of Wisconsin Department of Family Medicine and Community Health. We track daily totals of absentee counts in each category. In addition, we calculate the average daily count for each week of the school year in each category. The data flow is illustrated in Figure 3.

**Figure 3.**
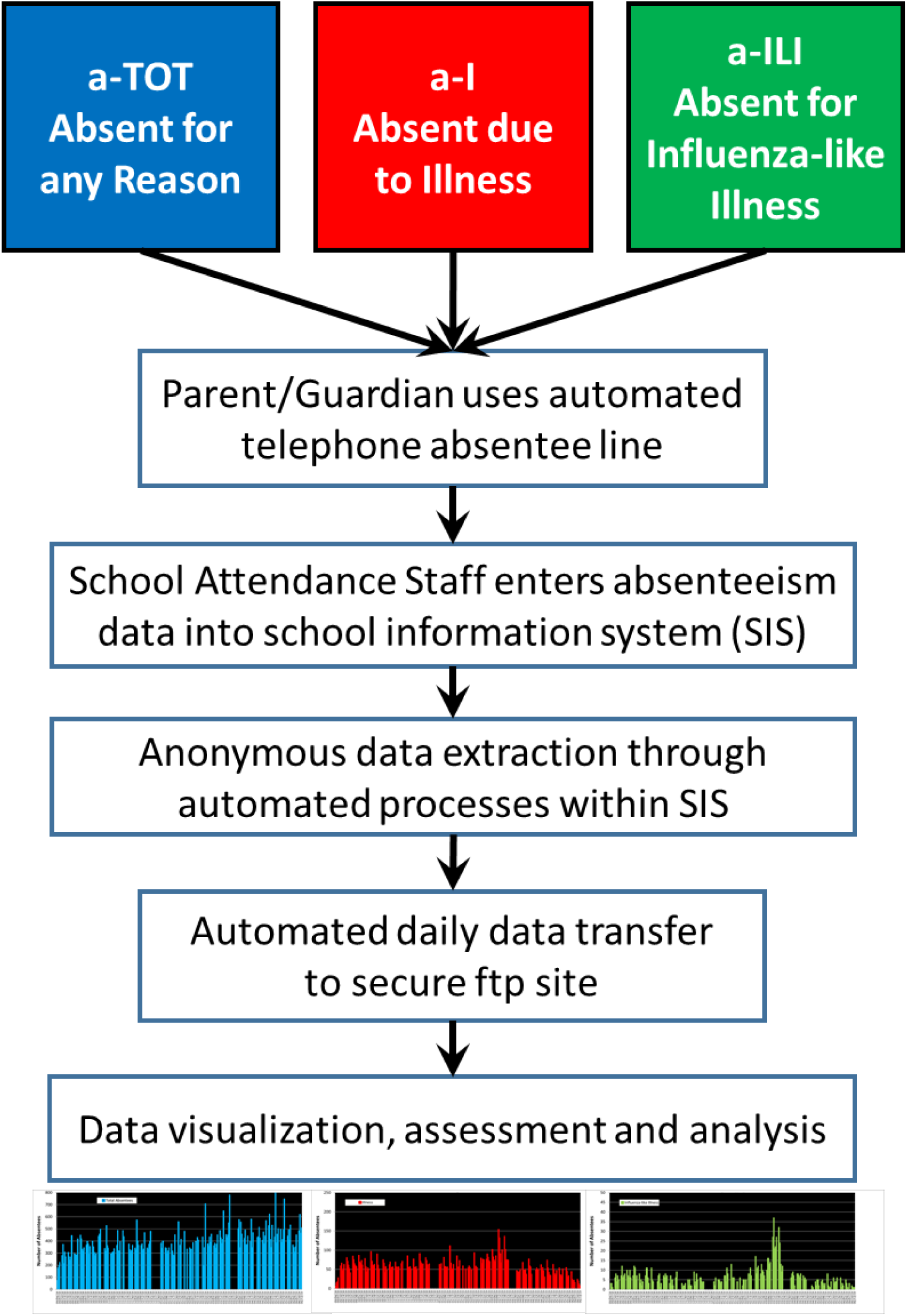
Flow diagram of absenteeism data from telephone reporting by parents/guardians, to entry into the student information system at the Oregon School District, to data transfer to the ORCHARDS research team.

#### School incentive

Each school receives $4,000 per year to defray costs associated with IT support and effort by the attendance staff.

### IVc. Assessment of influenza and absenteeism in children through home visits

#### Contact and screening

To comply with FERPA, interested parents/guardians voluntarily call the study line to determine if their child qualifies for a home visit. Students are not required to be absent to participate and it is not necessary for school to be in session for home visits to occur. If a student meets the inclusion criteria, a 20-minute home visit occurs within 2 days of initial phone contact and within 7 days of symptom onset.

#### Inclusion criteria include

1. student attends, or is eligible to attend (e.g., home schooled), a school within the OSD
2. student has an illness characterized by at least 2 of 6 acute respiratory infection (ARI)/ILI symptoms (nasal discharge; nasal congestion; sneezing; sore throat; cough; fever)
3. student scores at least 2 points on the Jackson scale (26-28)

#### Exclusion criteria include

1. household member listed on Wisconsin Department of Corrections Sex Offender Registry (http://offender.doc.state.wi.us/public/) [for the safety of field staff]
2. illness onset more than 7 days before anticipated time of specimen collection
3. anatomical defect for which nasal specimen collection is contraindicated
4. student participated too recently (<7 days during peak influenza period and <30 days during other times, as determined by medically-attended surveillance program)

At the end of the screening process for a student meeting study criteria, family members are invited to participate in an optional household influenza transmission sub-study. Participation is allowed even if individual members opt out or are unable to complete the entire study.

#### Inclusion criteria include

1. individuals of any age/gender residing in the same household as ORCHARDS participant
2. fluent in English
3. able to provide appropriate consent/assent

#### Exclusion criteria include

1. anatomical defect for which nasal sampling is contraindicated
2. household participated too recently (<7 days during peak influenza period and <30 days during other times, as determined by medically-attended surveillance program)

#### Acquiring informed consent/assent

Research coordinators obtain written informed consent from parents/guardians and/or adult students, and assent from younger students using forms tailored to reading levels based on age. Consent allows assessment of immunization records through the Wisconsin Immunization Registry and advanced laboratory analyses of respiratory specimens. Participants can opt for study personnel to contact them about future research opportunities. The consent process changed on August 27, 2018 to allow recurrent visits throughout the full academic year.

#### Data collection

Demographic, epidemiologic, and symptom data (Table 2) are collected on an Acute Respiratory Infection and Influenza Surveillance Form. This form (Supplement 1) is based on an instrument used in clinical settings since 2009 as part of the Wisconsin component of the Influenza Incidence Surveillance Project (W-IISP).(8) Accordingly, the data collected are comparable to data from other surveillance systems.

**Table 2.** ORCHARDS data elements obtained at all home visits. For details on data collection form, see supplement 1.

| **Data Element** | **Description** |
| --- | --- |
| **Epidemiological** |  |
| Time from symptom onset | Number of days from illness onset to home visit |
| Exposure to similar illness | Exposure to similar illness 1-3 days prior to illness onset |
| Likely source | Classmate, friend, family member, other |
| Household members | Identification of all household members and presence of similar illness |
| Recent travel | Travel to destination $\geq 50$ miles in week before onset |
| Exposure to farm animals | Contact with farm animals prior to illness onset |
| **Demographic** |  |
| Age | years |
| Sex | male / female / other |
| School | Assigned school |
| Race | Standard categories |
| Ethnicity | Standard categories |
| **Clinical** |  |
| Severity | Mild, moderate, severe as reported by parent/guardian |
| Measured temperature | Temperature taken by ORCHARDS staff at home visit |
| Presence of symptoms | Presence/absence of 17 symptoms; other recorded |
| Use of antipyretics in last 6 hours | Reported by parent /guardian |
| Recent influenza antiviral use | Reported by parent /guardian |
| Receipt of current seasonal influenza vaccine | Reported by parent /guardian (verified through registry) |
| Medical visit prior to home visit | Reported by parent /guardian |
| Medical visit planned after home visit | Reported by parent /guardian |

#### Respiratory specimen collection and handling

A nasal swab specimen is collected using a Puritan^®^ Sterile Foam Tipped Applicator for same-day rapid influenza diagnostic testing (RIDT). Starting on September 4, 2019, we started collecting the nasal specimen from both nostrils. In addition, a nasopharygeal (NP) or high oropharyngeal specimen (OP) is obtained using a Copan FLOQSwabs™ flocked swab. The NP/OP swab is placed into a 3.0 ml Remel MicroTest™ M4RT^®^ Transport viral transport medium (VTM) tube, sealed and placed into a small resealable plastic biohazard bag, and maintained at 2-8°C. Following processing for RIDT, the residual nasal swab is added to the VTM containing the NP/OP swab. A courier delivers the VTM tube to the Wisconsin State Laboratory of Hygiene (WSLH), usually within 24 hours of collection.

#### Incentives and feedback

Student participants receive a $20 gift card at the end of the home visit. Research coordinators call parents/guardians with RIDT results within 24 hours (usually less than 4 hours) of the home visit. Laboratory confirmed results are mailed to families within 2 weeks of a home visit, and are accepted as documentation for an excused absence by the OSD.

### IVd. Within-household influenza transmission sub-study

#### Contact and screening

At the ORCHARDS home visit, families expressing interest in the sub-study receive a packet containing sub-study information, instructions, consents/assents, and a small collection kit for each household member. The collection kit contains a data form, 2 nasal swabs, and 2 small biohazard bags, each of which contain a 3.0 ml Remel MicroTest™ M4RT^®^ VTM tube. VTM tubes and swabs for Day 0 and Day 7 are individually marked.

The research coordinator explains each component of the family packet to all household members who are present and reviews consent/assent forms. A designated adult assures that all interested household participants sign the consent/assent forms prior to specimen collection. Household members are responsible for collecting their own specimens on Day 0 (within 24 hours of home-visit) and Day 7 (seven days after the initial collection). The timing (Day 7) for follow-up data and specimen collection is based upon prior studies of household transmission of influenza.(29-31)

#### Data collection

The ORCHARDS Household Study Form (Supplement 2) is used to collect data pertaining to the following:

1. Household composition: the number and characteristics of people sharing the household of an ORCHARDS participant. Information includes relationship to the ORCHARDS participant, age, gender, number of bedrooms in home, employment outside of home, school attendance, and daycare attendance.
2. Household member illness assessment on Day 0 (“Today” section): influenza vaccine status for the current season, and information about any cold or flu-like illnesses occurring over the past 7 days (Table 3).
3. Household member illness assessment on Day 7, one week after the initial visit (“Follow-up” section): information about any cold or flu-like illnesses occurring over the past 7 days (Table 3).

**Table 3.** ORCHARDS household transmission study data elements obtained from all household participants. Some data elements are collected on Day 0 only, while other are collected on Day 0 and Day 7. For details on data collection form, see supplement 2.

| Data Element | Description | Response on |  |
| --- | --- | --- | --- |
|  |  | Day 0 | Day 7 |
| Epidemiological |  |  |  |
| Number of bedrooms in home | Measure of house size | x |  |
| Work outside of home | For adult household members (y/n) | x |  |
| School attendance | For school aged household members (y/n) | x |  |
| Daycare attendance | For younger children (y/n) | x |  |
| Presence of respiratory illness or symptoms in the past 7 days | Assessment for incidence of respiratory infections (y/n) | x | x |
| Time of onset of symptoms | Assessment of onset date | x | x |
| Demographic |  |  |  |
| Age | years | x |  |
| Gender | male / female / other | x |  |
| Relationship to ORCHARDS student | parent, guardian, sibling, grandparent, other | x |  |
| Clinical |  |  |  |
| Severity of illness | Mild, moderate, or severe as reported by parent/guardian | x | x |
| Presence of symptoms | Presence/absence of 10 symptoms | x | x |
| Receipt of current seasonal influenza vaccine | Reported by parent /guardian (y/n) (verified through immunization registry) | x |  |
| Medical visit for this illness | Reported by parent /guardian (y/n) | x | x |
| Venue of medical visit | Clinic, urgent care, emergency department | x | x |
| Diagnosis from medical visit | Reported by parent /guardian | x | x |
| Antibiotic or antiviral prescribed | Reported by parent /guardian | x | x |
| Sent to hospital | Reported by parent /guardian (y/n) | x | x |
| Absent from work/ school for illness | Number of absentee days (y/n) | x | x |
| Number of days absent | Number of absentee days | x | x |

#### Respiratory specimen collection and handling

Research coordinators instruct household members on respiratory specimen collection from one nostril using a Copan FLOQSwabs™ flocked mid-turbinate swab (September 6, 2016 through October 15, 2017) or an anterior nasal Puritan^®^ Sterile Foam Tipped Applicator (starting on October 16, 2017). On September 4, 2019, participants were instructed to swab both nostrils with a single swab. Each household participant collects the specimen without staff observation the day of the home visit (Day 0) and again seven days later (Day 7). Contact is made prior to or on Day 7 to reinforce completion of sub-study forms and specimens collection. When retrieving household collection kits, research coordinators review forms for completion and ensure specimens were collected and correspond with the appropriate labels.

#### Household incentive for participation

As an incentive for completing the household sub-study, participating families receive a $50 gift card to local businesses when specimens are retrieved.

### IVe. Laboratory procedures

#### Rapid influenza diagnostic test

We use the Quidel^®^ Sofia^®^ Influenza A+B fluorescent immunoassay for initial assessment of nasal specimens for ORCHARDS participants.(32) RIDT is performed at a nearby clinical facility within 6 hours of specimen collection following instructions outlined in the package insert. RIDT is not performed on specimens from household members.

#### Influenza rRT-PCR

All specimens from students and household members (Day 0 and Day 7) are tested at WSLH for influenza A and B virus and Human RNase P (RP) using the in-vitro diagnostic (IVD) FDA-approved CDC Human Influenza Virus Real-time RT-PCR Diagnostic Panel (Cat.# FluiVD03) (33). Per protocol, the detection of RP, an indicator of human epithelial cells, with a cycle threshold (Ct) value <38 determines specimen adequacy for influenza PCR testing. Influenza A and B positive specimens are subtyped using the same testing kit.

#### Multipathogen testing

All specimens from ORCHARDS students are tested for influenza A, influenza B, rhinovirus/enterovirus, adenovirus, parainfluenza virus (1, 2, 3, 4), coronavirus (HKU1, NL63, 229E, OC43), respiratory syncytial virus (A, B), human metapneumovirus and human bocavirus, and 2 atypical bacterial pathogens, *Chlamydia pneumoniae* and *Mycoplasma pneumoniae*, using the multiplexed PCR respiratory pathogen panel (RPP: Luminex NxTAG Respiratory Pathogen Panel).(34)

#### Whole genome sequencing

For a subset of influenza-positive subjects, we conduct next-generation genome sequencing at the WSLH using the Illumina MiSeq™ platform. Sequence reads are trimmed using Trimmomatic with a 4 bp sliding window quality score cutoff of Q30.(35) The trimmed sequence reads are mapped against vaccine strain HA reference sequences using Geneious version 11.0.5.(36)

#### Quality control

The influenza rRT-PCR and multipathogen testing at the WSLH goes through a rigorous validation process to meet federal Clinical Laboratory Improvement Amendments licensing standards where sensitivity, reproducibility, accuracy and specificity have been demonstrated.

#### Specimen archiving

Aliquots of residual specimens, including all positive, negative, and nucleic acid are archived at ≤-70°C at WSLH indefinitely or until CDC requests shipment. When requested, residual specimens and nucleic acid are submitted to CDC on dry ice by overnight shipment according to current practice and CDC requests.

### IVf. Validation of influenza vaccination

We validate influenza immunization status for all ORCHARDS students using the Wisconsin Immunization Registry (WIR).(37) Students are determined to be fully vaccinated or not at the time of specimen collection using the guidance provided by the US Advisory Committee on Immunization Practices.(38) Subjects also provide consent to evaluate receipt status of other vaccines within WIR.

### IVg. Data security and integrity

All identifying information on subjects is secured within UW Health using a separate password-protected security REDCap (Research Electronic Data Capture) database.(39) All other project data are kept in password-protected security-ensured REDCap databases. Data analysts are restricted to viewing outcome data, and are unable to access personal identifiers.

### IVh. Community engagement and study promotion

The primary mode of ORCHARDS recruitment is the reminder within the absenteeism reporting system. Information is also provided at the OSD’s unified registration each August and through e-mails to OSD families. We employ flyers, posters, and brochures at community sites, presentations at community events, mailings, postcards, the study website (www.fammed.wisc.edu/orchards/), and Facebook page (www.facebook.com/orchardstudy/). Moreover, the study team has extensive personal connections with the community through long-term residency within the OSD, children attending school in the district, and participation in school-based activities.

### IVi. IRB and project oversight

All components of this proposed study were reviewed and approved by the Human Subjects Committees of the Education and Social/Behavioral Sciences IRB (initial approval on September 4, 2013) and the University of Wisconsin Health Sciences-IRB (initial approval on December 5, 2013, with additional approvals as the protocol expanded and modified). The study is in full compliance with the Health Insurance Portability and Accountability Act of 1996 (HIPAA), FERPA, and all other federally mandated human subjects regulations. The US Office of Management and Budget has approved all forms used in this study.

### IVg. Reference data: assessment of influenza in the surrounding community

A fully independent and long-standing influenza surveillance system is available to assess MAI in and around the OSD. W-IISP is sponsored by CDC and the Council of State and Territorial Epidemiologists and has conducted prospective, active surveillance for influenza and other respiratory viruses in medically attended patients with ARI or ILI since October 2009 (8,40). The ORCHARDS research team organizes this surveillance system. Active surveillance occurs at 5 family practice clinics within or adjoining the OSD. Specific clinic locations include Belleville, Oregon, Madison (2 sites) and Verona (Figure 2). Together, the clinics recorded 412,752 ambulatory visits between June 29, 2014 and June 29, 2019 and assessed 4,069 individual ARI patients.

NP/OP swabs are collected from a subset of ambulatory patients with ARI and ILI. All specimens are tested at the WSLH for influenza A and B using the rRT-PCR (33) and, for other viruses, using a multiplexed PCR RPP.(34) Virological characterization is available for all (n = 4,069) individual patient visits.

## V. Results

### Va. Absenteeism data

Over the first 5 years of ORCHARDS, we have evaluated 3,260,461 student days (enrollment multiplied by school days). Total absenteeism accounted for 301,427 (9.2%) of potential student days, a-I accounted for 58,126 (1.8%) student days, and a-ILI accounted for 6,634 (0.2%) student days. The daily counts for each type of absenteeism, showing the variability, are depicted in Figure 4. The annual levels of absenteeism were similar across all 5 years (Table 4).

**Table 4.** Enrollment estimates and absenteeism by defined type for the Oregon School District, Oregon, Wisconsin: 2014—2019.

|  | **2014-2015** | **2015-2016** | **2016-2017** | **2017-2018** | **2018-2019** |
| --- | --- | --- | --- | --- | --- |
| Total Estimated Enrollment* | 3,588 | 3,713 | 3,749 | 3,828 | 3,867 |
| Total Curricular Days | 176 | 177 | 175 | 176 | 174 |
| Total Possible Student Days | 629,024 | 635,076 | 649,775 | 673,728 | 672,858 |
| a-TOT Days<br>(% of total) | 56,477<br>(8.98%) | 55,871<br>(8.80%) | 56,511<br>(8.70%) | 63,476<br>(9.42%) | 69,092<br>(10.3%) |
| Average Daily a-Tot<br>(std. dev.) | 320.9<br>(78.4) | 315.7<br>(79.2) | 322.9<br>(80.4) | 360.7<br>(96.3) | 397.1<br>(106.2) |
| a-I Days<br>(% of total) | 12,024<br>(1.91%) | 12,158<br>(1.91%) | 11,922<br>(1.83%) | 11,710<br>(1.74%) | 10,312<br>(1.53%) |
| Average Daily a-I<br>(std. dev.) | 68.3<br>(39.0) | 68.7<br>(27.0) | 68.1<br>(27.0) | 66.5<br>(30.1) | 59.3<br>(21.7) |
| a-ILI Days<br>(% of total) | 1,314<br>(0.21%) | 1,609<br>(0.25%) | 1,309<br>(0.20%) | 1,261<br>(0.19%) | 1,141<br>(0.17%) |
| Average Daily a-ILI<br>(std. dev.) | 7.5<br>(7.3) | 9.1<br>(6.6) | 7.5<br>(5.9) | 7.2<br>(5.8) | 6.6<br>(5.5) |

\* Total estimated enrollment excluding 4K students (for whom absenteeism data is not submitted). Data extracted from: Wisconsin Department of Public Instruction, Wisconsin Information System for Education data Dashboard: https://wisedash.dpi.wi.gov/Dashboard/portalHome.jsp

**Figure 4.**
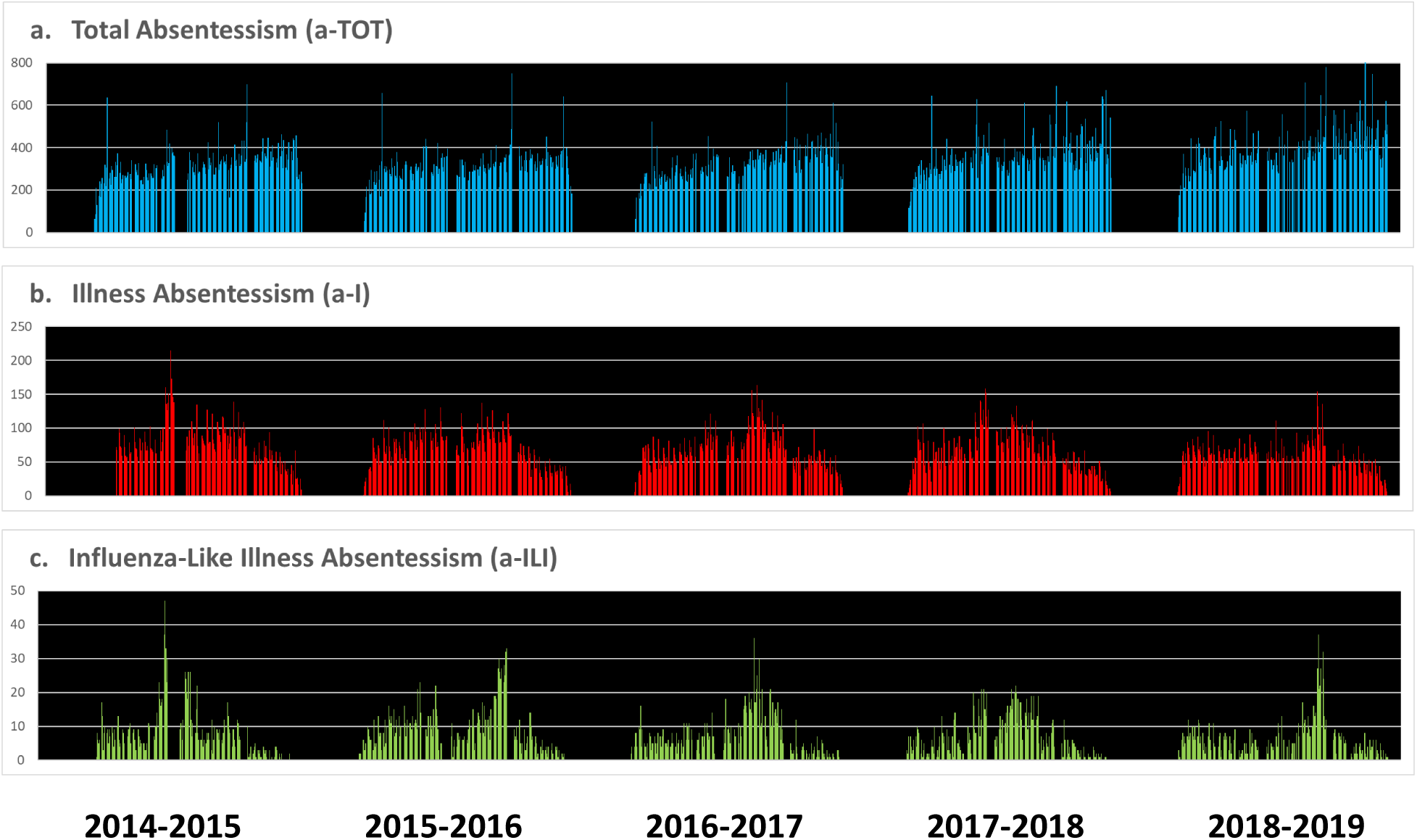
Daily counts of all absent students (a-TOT: panel a), students for whom an illness is reported (a-I: panel b) and students absent with influenza-like illness (a-ILI: panel c) occurring over five consecutive school years at the Oregon school District, Wisconsin, USA from September 2014 through June 2019.

### Vb. Home visits and basic virology

We completed 1,728 home visits for children with ARI. Children ranged in age from 4 to 18 years (mean ± std.dev. = 9.9±2.5 years). There were more male (57%) than female (43%) participants. Home visits occurred, on average, 56.3±46.5 hours after onset of symptoms. The number of home visits per day were positively correlated with a-Tot (r_s_=0.20; p<0.001), a-I (r_s_=0.41; p<0.001) and a-ILI (r_s_=0.40; p<0.001). Most children (79%) reported school absenteeism due to the current illness episode.

Pathogens were detected in 1,115 (65%) specimens; the majority (99%) of these were viral. Co-detections of viruses were noted in 66 students (6% of individuals with virus detections). Influenza was the most commonly detected virus, noted in 402 (23%) students, followed by rhinovirus/enterovirus. The numbers of viruses detected are provided in Table 5.

**Table 5:** Detections of viruses from ORCHARDS participants through home visits (January 5, 2015 to June 30, 2019). The counts include dual detections (n=64) and triple detections (n=2). Not shown are data for influenza A (unable to subtype: n=3), *Mycoplasma pneumoniae* (n=12), and *Chlamydia pneumoniae* (n=1). Percentages are expressed as detections of specified virus groups divided by all detections.

|  | Respiratory Virus Detected |  |  |  |  |  |  |  |  |
| --- | --- | --- | --- | --- | --- | --- | --- | --- | --- |
|  | Ad | CoV | FluA(H1) | FluA(H3) | FluB | hMPV | PIV | R/E | RSV |
| Number of Detections | 32 | 169 | 77 | 224 | 98 | 74 | 77 | 368 | 47 |
| Percent | 2.7 | 14.3 | 6.5 | 18.9 | 8.3 | 6.3 | 6.5 | 31.1 | 3.9 |

**Ad**=adenovirus; **CoV**=coronavirus; **Flu**=influenza virus; **RSV**=respiratory syncytial virus; **hMPV**=human metapneumovirus; **PIV**=parainfluenza virus; **R/E**=rhinovirus/enterovirus

### Vc. Validation of a-ILI definition

A simple, validated definition of influenza-like illness–related absence (a-ILI) is a prerequisite for monitoring. We used multivariate binary logistic regression to assess the relationships between pathogens and a-ILI within individual students while adjusting for age and sex. Cases for which no viruses were identified served as the reference set. Of the participating students evaluated between January 5, 2015 and June 30, 2019, 1,552 (90%) with ARI onset while school was in session remained in the analysis. Influenza [FluA(H1), FluA(H3), FluB], and adenovirus (Ad) were significantly associated with positive likelihoods of being absent with an ILI; the presence of rhinovirus/enterovirus (R/E) was associated with a negative likelihood of a-ILI (Table 6). Other viruses were not associated with a-ILI. Accordingly, absence from school due to ILI using a simple definition (absent with fever and a respiratory symptom) is strongly associated with laboratory-confirmed influenza.

**Table 6:**
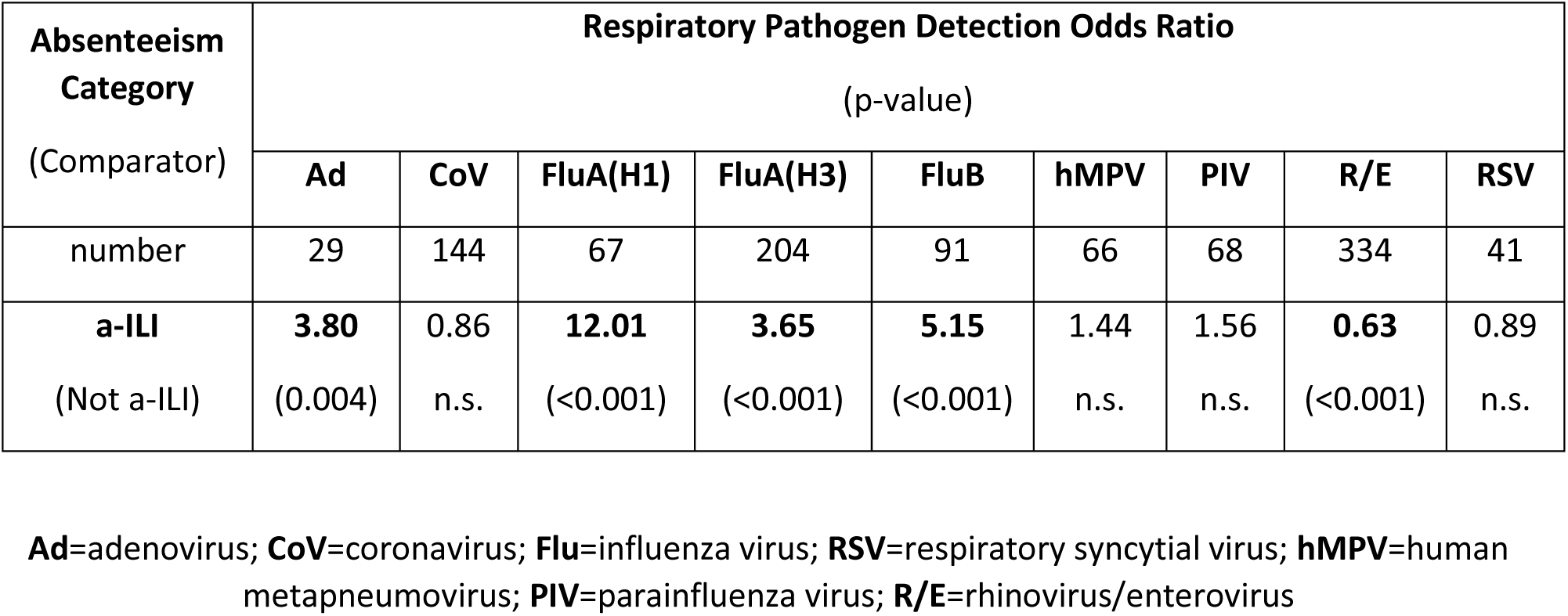
Associations (odds ratios) of specific respiratory viruses with absences from school due to influenza-like illness (a-ILI) for K-12 children in the Oregon School District, Wisconsin, USA (January 5, 2015, to June 30, 2019).

## VI. Discussion

In contrast to routine national surveillance systems that rely on sentinel outpatient or inpatient data on medically attended cases of ILI, ORCHARDS uses a community-based design. It is well recognized that most cases of influenza do not present for medical attendance (40). Even less common are hospitalized cases of influenza and influenza-related mortality (41-42). It is also recognized that the attack rates of influenza are much higher in school-aged children than for any other demographic group (40), and that influenza has a significant contribution to school absenteeism.(43) Accordingly, ORCHARDS was designed to detect and evaluate—over multiple seasons—the temporal trends of influenza detection among the age group which is central to community-wide influenza transmission (school-aged children), and which may be less represented among MAI cases.

A number of studies evaluating absenteeism and influenza predate ORCHARDS (15-18, 44), but many have been limited by evaluating a single outbreak. Influenza does not follow a regular pattern in an area but rather encompasses outbreaks of variable magnitudes and temporal patterns due to differing influenza types, subtypes and clades (45,46); this necessitates multiyear assessment of monitoring systems to assess generalizability. Moreover, influenza outbreaks can occur at any time over a fairly wide seasonal range (47), thus making assessments over several seasons necessary to evaluate for effects of timing. Finally, an observational approach allows accumulation of multiple periods of planned and unplanned (weather-related) school breaks that may allow evaluation of school closure for outbreak response.

ORCHARDS takes advantage of a pre-existing, long-standing, and highly effective influenza surveillance system that has provided consistent evaluation of seasonal and pandemic influenza since October 2009 as a “gold standard” for daily comparability. This parallel system is based on MAI surveillance at 5 primary care clinics which overlap with the study catchment area and uses very similar data instruments and identical laboratory methods.

## VII. Conclusions

The community involvement, longitudinal nature, and external comparability make ORCHARDS a unique study platform to evaluate the role of school-aged children on influenza transmission and the utility of cause-specific absenteeism monitoring for identifying influenza outbreaks. Initial findings suggest the study design is succeeding in collecting appropriate data to achieve study objectives. Early detection of influenza within a community is critical for public health mitigation and the reduction of overall disease burden. Additional studies are needed to determine the effect of school closure on influenza transmission.

## Data Availability

The datasets generated and/or analyzed during the current study are not publicly available because the study is ongoing, but may be available from the corresponding author on reasonable request.

## Abbreviations

4k: 4-year-old pre-kindergarten
a-I: absenteeism due to illness
a-ILI: absenteeism due to influenza-like illness
a-TOT: all-cause absenteeism
ARI: acute respiratory infection
CDC: Centers for Disease Control and Prevention
CoV: coefficient of variation
FERPA: Family Educational Rights and Privacy Act
Flu: influenza
FDA: Food and Drug Administration
HIPPA: Health Insurance Portability and Accountability Act
ILI: influenza-like illness
IT: information technology
IRB: institutional review board
MAI: medically attended influenza
NP: nasopharyngeal
ORCHARDS: Oregon Child Absenteeism Due to Respiratory Disease Study
OSD: Oregon School District
OP: oropharyngeal
PCR: polymerase chain reaction
RIDT: rapid influenza diagnostic test
RedCap: Research Electronic Data Capture
RPP: respiratory pathogen panel
R/E: rhinovirus/enterovirus
RP: human RNAse P
SIS: school information system
W-IISP: Wisconsin Influenza Incidence Surveillance Project
WIR: Wisconsin Immunization Registry
WSLH: Wisconsin State Laboratory of Hygiene
VTM: viral transport medium

## Acknowledgements

We are indebted to Dr. Brian Bussler and Dr. Leslie Bergstrom (former and current OSD Superintendent), the OSD Board of Education, Mr. Jon Tanner (IT Director), and many individuals in the attendance and health offices for project facilitation, and to hundreds of students and their families within the OSD for enthusiastic participation in ORCHARDS. The Wisconsin State Lab of Hygiene provided excellent laboratory support and advice on best practices.

