## Supplemental 1 for "The Oregon Child Absenteeism Due to Respiratory Disease Study (ORCHARDS): Rationale, Objectives, and Design"

U.S. Department of Health and Human Services (HHS)
Centers for Disease Control and Prevention (CDC)

OMB Approved
Control # 0920-1039

ORegon CHild Absenteeism due to Respiratory Disease Study

### Acute Respiratory Infection and Influenza Surveillance Form

Criteria for patient selection and testing
- symptom onset within 4 days
- any two of the following
  - rhinorrhea
  - nasal congestion
  - sneezing
  - sore throat
  - cough
  - fever

**School ID: 4K P N B R M H**

**Participant ID:** ____________

**Age:** ________________

**Date of Collection:** _____________

- - - - - - - - - - - - - - - - - - - - - - - - - - - - - - - - - - - - - - - - -

**Days between illness onset and today's visit:** ____ days

**Exposure to a similar illness 1-3 days prior to ARI onset?** Yes No

**Likely Source:** Classmate Friend Family Member (Adult / Sibling) Other: __________________

**Household Member** (circle if living in household, check box if ill with similar ARI):

| | | | |
|---|---|---|---|
| Grandmother ☐ | Grandfather ☐ | Mother (*Female Guardian)* ☐ | Father (*Male Guardian)* ☐ |
| O/Y Sibling #1 ☐ | O/Y Sibling #2 ☐ | O/Y Sibling #3 ☐ | Other Adult:________ ☐ |
| O/Y Sibling #4 ☐ | O/Y Sibling #5 ☐ | O/Y Sibling #6 ☐ | Other Child:________ ☐ |

**Recent Travel?** Yes No **Recent Exposure to Farm Animals?** Yes No

**Severity of Illness** (circle)**:** Mild Moderate Severe

**Race:** White Am Indian or Ak Native Asian Black Native Hawaiian or Other Pacific Islander

**Ethnicity:** Hispanic Non-Hispanic

**Measured Temperature** ___ $^{o}$F **Antipyretic use within the last 6 hours?** Yes No

**Symptoms** (circle all that are present)**:**

| | | | | | |
|---|---|---|---|---|---|
| Fever | Chills | Cough | Wheezing | Runny Nose | Sore Throat |
| Malaise | Myalgia | Arthralgia | Nasal Congestion | Headache | Ear Pain |
| Anorexia | Vomiting | Abdominal Pain | Diarrhea | Conjunctivitis | Other: ____________ |

**Influenza antiviral treatment for this illness prior to this visit?** Yes No

**Seasonal influenza vaccine prior to this illness?** Yes No

**Visit to health care provider for this illness prior to home visit?** Yes (specify when _________________) No

**Visit to health care provider planned in next few days?** Yes (specify when _________________) No

Indicate Specimen Type(s) for PCR testing: nasopharynx posterior pharynx

- - - - - - - - - - **ITEMS BELOW THIS LINE FOR LABORATORY ONLY** - - -

**Sofia Result:** ☐ **flu A** ☐ **flu B** ☐ **both** ☐ **negative** ☐ **invalid**

**Sample Code:** _ORCHARDS_ - _____ - ____ - ______
site week staff sample

_________________________________________________________________________

Public reporting burden of this collection of information is estimated to average 30 minutes per response, including the time for reviewing instructions and completing and reviewing the collection of information. An agency may not conduct or sponsor this survey, and a person is not required to respond to a collection of information, unless it displays a currently valid OMB Control Number. Send comments regarding this burden estimate or any other aspect of this collection of information, including suggestions for reducing this burden to CDC/ATSDR Reports Clearance Officer, 1600 Clifton Road NE, MS D-74, Atlanta, Georgia 30333; ATTN: PRA 0920-1039.
