## Supplemental 2 for "The Oregon Child Absenteeism Due to Respiratory Disease Study (ORCHARDS): Rationale, Objectives, and Design"

### ORCHARDS HOUSEHOLD STUDY FORM

**School ID:** _______________

**Participant ID:** ____________

**NASAL SWAB**

**ID**

**HOUSEHOLD MEMBER NAME:** ______________________

**RELATIONSHIP TO STUDENT:** _______________________

**Age:** _____ **Do you work outside the home?** Yes No **Number of bedrooms:** _____

**Gender:** F M **Do you attend school?** Yes No **Do you attend Daycare?** Yes No

- - - - - - - - - - - - - - - - - - - - - - - - - - - - - - - - - - - - - - - - - - - - - - - - -

**TODAY** Day 0 ( ___/___/___ )

**-Did you receive an influenza vaccine this year (after August 1, 2019)?** Yes No

**Have you had cold or flu-like symptoms in the past 7 days?** Yes No (if **No**, then you are done until next week)

**If yes: How many days ago did your symptoms start?** _______

**How severe are/were your symptoms?** Mild Moderate Severe

**What symptoms have you had in the past 7 days?** (circle all that have been present)

| | | | | |
|---|---|---|---|---|
| Fever | Chills | Cough | Runny Nose | Sore Throat |
| Tiredness | Body Aches | Headache | Poor Appetite | Nasal Congestion |

**Were you seen by a healthcare provider?** Yes No **Where?** Usual Clinic Urgent Care ER

What diagnosis were your given? ______________________

Were you given an antibiotic or antiviral medication? Yes No ________________________

Were you sent to the hospital? Yes No

**Did you miss school or Work?** Yes No **If yes, how many days did you miss?** ______

- - - - - - - - - - - - - - - - - - - - - - - - - - - - - - - - - - - - - - - - - - - - - - - - -

**FOLLOW-UP** Day 7 ( ___/___/___ )

**Have you had cold or flu-like symptoms in the past 7 days (since our previous visit)?** Yes No

**If yes: How many days ago did your symptoms start?** _______

**How severe are/were your symptoms?** Mild Moderate Severe

**What symptoms have you had in the past 7 days?** (circle all that have been present)

| | | | | |
|---|---|---|---|---|
| Fever | Chills | Cough | Runny Nose | Sore Throat |
| Tiredness | Body Aches | Headache | Poor Appetite | Nasal Congestion |

**Were you seen by a healthcare provider?** Yes No **Where?** Usual Clinic Urgent Care ER

What diagnosis were your given? ______________________

Were you given an antibiotic or antiviral medication? Yes No ________________________

Were you sent to the hospital? Yes No

**Did you miss school or Work?** Yes No **If yes, how many days did you miss?** ______
