## Supplemental 3 for "The Oregon Child Absenteeism Due to Respiratory Disease Study (ORCHARDS): Rationale, Objectives, and Design"

**Oregon School District Infinite Campus Data Export Code:**

```
<?php


require_once 'ms_quick_db.php';


$todays_date = date('Y-m-d');

$filename = $todays_date.'.csv';


$query = "select distinct a.personID,a.calendarID,case en.grade when 'KG' then '0' else
en.grade end as mygrade,e.code

from attendance a

inner join attendanceExcuse e on a.excuseID = e.excuseID

inner join enrollment en on a.personID = en.personID and a.calendarID = en.calendarID

inner join calendar c on a.calendarID = c.calendarID

where a.date = '$todays_date' and e.status = 'A'

order by a.calendarID,mygrade,a.personID";


/* quick test to see what schools ids are available

$query = "select * from calendar where YEAR(endDate) = '2017'";

*/


/*

echo "Using query:\n";

echo "=========================================\n";

echo "\n\n";
```

```
echo $query;

echo "\n\n";

echo "========================================\n";

echo "\n\n";

*/


//UPDATE THIS EACH YEAR, calendarID => School

$schools = array(

'145' => 'Netherwood Elementary',

'144' => 'Brooklyn Elementary',

'146' => 'Prairie View Elementary',

'148' => 'Rome Corners Intermediate',

'147' => 'Oregon Middle School',

'143' => 'Oregon High School');



//seed output arrays with some base data

$data = array();


//echo "Processing $filename\n";

$res = ms_quick_rows($query);


/*

foreach ($res as $r) {
```

```
print_r($r);
}

die();
*/

//personID, calendarID, grade, code
foreach($res as $r) {
  $personID = $r[0];
  $calendarID = $r[1];
  $grade = intval($r[2]);
  $code = $r[3];

  if($code == 'AILI' || $code == 'AI') {
    if(isset($data[$calendarID][$grade]['ai'])) {
      $data[$calendarID][$grade]['ai'] += 1;
    } else {
      $data[$calendarID][$grade]['ai'] = 1;
    }
  }

  if($code == 'AILI') {
    if(isset($data[$calendarID][$grade]['ili'])) {
      $data[$calendarID][$grade]['ili'] += 1;
```

```
    } else {
      $data[$calendarID][$grade]['ili'] = 1;
    }
  }

  if(isset($data[$calendarID][$grade]['total'])) {
    $data[$calendarID][$grade]['total'] += 1;
  } else {
    $data[$calendarID][$grade]['total'] = 1;
  }
}

$output = array();

foreach($data as $calID => $ar) {
//  echo "Processing CalendarID $calID\n";
  foreach($ar as $gd => $x) {
    if(isset($x['ili'])) {
      $ili = $x['ili'];
    } else {
      $ili = 0;
    }
    if(isset($x['ai'])) {
      $ai = $x['ai'];
```

```
    } else {
      $ai = 0;
    }
    //echo $schools[$calID].', '.$todays_date.', '.$gd.', '.$ili.', '.$x['total']."\n";
    $output[] = array($schools[$calID],$todays_date,$gd,$ili,$ai,$x['total']);
  }
}

$fp = fopen('/var/www/orchards/export/'.$filename,'w');

$headers = array('School','Date','Grade','ILI','AI','Total');

fputcsv($fp, $headers);
foreach($output as $o) {
  fputcsv($fp, $o);
}
fclose($fp);
```
